# Wastewater Treatment Attenuates Ecological and Human Health Resistome Risk

**DOI:** 10.64898/2026.09.12.26362900

**Authors:** Rose O. Elliott, Susheel Bhanu Busi, Lindsay K. Newbold, Michael J. Bowes, Linda K. Armstrong, David J. E. Nicholls, Hyun S. Gweon, Barbara Kasprzyk-Hordern, Daniel S. Read, Holly Tipper

## Abstract

Wastewater treatment is an important barrier to the environmental dissemination of antimicrobial resistance (AMR). However, its effectiveness is typically assessed using measures that do not directly capture resistance gene mobility or pathogen association. Here, we applied a configuration-based genomic risk framework (MetaCompare2.0) to 100 metagenomes (50 paired influent and effluent samples) collected from nine wastewater treatment works over 11 months, quantifying resistome risk based on associations between antimicrobial resistance genes (ARGs), mobile genetic elements, and host taxa. Influent samples exhibited highly variable ecological risk associated with mobile ARGs. Pathogen-associated human health risk was lower and more constrained. Wastewater treatment consistently reduced both ecological and human health risks, driving convergence towards uniformly lower risk resistome configurations. Risk attenuation was reproducible across treatment technologies and could not be explained by assembly quality or physicochemical variation. These findings indicate that wastewater treatment disrupts high-risk ARG configurations and highlight configuration-based genomic risk metrics as scalable tools for benchmarking environmental AMR mitigation.

## Introduction

Antimicrobial resistance (AMR) is widely recognised as one of the greatest biological and public health challenges of the twenty-first century. The emergence and dissemination of antimicrobial-resistant organisms compromise the effectiveness of modern medicine, threaten food security, destabilise healthcare systems, and impose growing burdens across human, animal, and environmental health sectors^1^. AMR is mediated by antimicrobial resistance genes (ARGs), which enable microorganisms to survive exposure to antimicrobial compounds and can disseminate between bacterial hosts via horizontal gene transfer (HGT) mediated by mobile genetic elements (MGEs) such as plasmids, transposons, and integrons^2^. Since the widespread introduction of anthropogenic antimicrobials, the mobilisation of ARGs from ancient environmental reservoirs into clinically relevant bacteria and transmissible MGEs has accelerated dramatically, reshaping microbial evolutionary trajectories on a global scale^3,4^. Consequently, AMR is no longer viewed solely as a clinical phenomenon, but increasingly as an ecological and evolutionary process emerging from interactions among microbial communities, selective pressures, environmental connectivity, and anthropogenic activity^5^.

Many resistance determinants predate the clinical antibiotic era and occur naturally within environmental microbial communities, where they fulfil diverse physiological and ecological functions^6^. The broader resistome therefore encompasses not only clinically recognised ARGs, but also intrinsic resistance determinants, environmental proto-resistance genes, latent resistance functions, and resistance-associated elements present across pathogenic and non-pathogenic microorganisms alike^7–9^. Importantly, resistance is not a fixed property of individual genes. Instead, it is an emergent feature shaped by genomic context, host background, regulatory state, ecological selection, and evolutionary opportunity^10^. ARGs that are currently environmentally benign may acquire clinical relevance following mobilisation into transmissible genetic platforms or clinically adapted hosts^10^.

Metagenomic sequencing has transformed the study of AMR by enabling culture-independent characterisation of resistomes across clinical, host-associated, and environmental systems^11^. Resistomes (the collection of all ARGs in a community^8^) are typically quantified by ARG richness, representing the diversity of resistance determinants detected, and ARG abundance, representing the proportion of sequencing reads assigned to resistance-associated functions^12^. As a result, most environmental AMR surveillance strategies rely heavily on abundance-based endpoints, commonly through quantitative polymerase chain reaction, metagenomic read mapping, or normalisation to bacterial marker genes such as 16S rRNA^13^. While these approaches provide valuable information about resistome composition and burden, they remain fundamentally limited as proxies for epidemiological risk because ARG abundance alone does not resolve dissemination potential, host range, mobility, persistence, or the likelihood of contributing to clinically consequential transmission events^14,15^.

Environmental AMR surveillance faces a fundamental conceptual challenge. Resistance genes do not behave like conventional chemical pollutants whose risks often scale predictably with concentration^16^. So, unlike classical microbial hazards, exposure to ARGs or resistant bacteria rarely produces immediate, observable health outcomes, but instead contributes to a “silent pandemic” in which adverse effects emerge only after ecological transmission, host colonisation, infection, and treatment failure^17^. In environmental matrices, ARGs can persist for long periods of time, transition among multiple hosts, and disseminate through complex microbial behaviours governing HGT, ecological selection, and interaction networks^18–21^, making their epidemiological significance context dependent and shaped by mobility potential, host association, persistence, selective pressures, and opportunities for onward transmission. Recognition of these limitations has driven a shift toward risk-oriented frameworks such as the “resistome risk” concept, which emphasises that ARGs linked to clinically important antibiotics, MGEs, and human pathogens pose disproportionately greater public health concern^22^. Importantly, in this context, “risk” does not represent a direct estimate of adverse health outcomes or transmission probability. Rather, it reflects the inferred epidemiological relevance of ARGs based on characteristics such as mobility, pathogen association, persistence, and opportunities for onward dissemination. Further work suggests that comparing resistome risk across environments may help identify ecological hotspots for ARG mobilisation and pathogen acquisition that warrant targeted mitigation^23,24^. This shift is crucial because environmental surveillance lacks the direct clinical endpoints available in healthcare settings, making translation of genomic data into quantitative public health relevance exceptionally challenging^25,26^, yet increasingly urgent, given evidence that environmentally mediated AMR transmission may contribute more substantially to clinical resistance burdens than previously recognised^21^.

Wastewater treatment works (WwTWs) occupy an important position within the environmental AMR continuum^27^. By aggregating municipal, hospital, industrial, and commercial waste streams, WwTWs concentrate microbial biomass, ARGs, MGEs, antimicrobial compounds, pathogens, nutrients, and diverse chemical stressors into interconnected ecological systems characterised by intense microbial interactions and selection pressure^27,28^. These environments may facilitate both the persistence and dissemination of AMR through enhanced opportunities for bacterial interactions and HGT^2^. Conventional treatment processes generally reduce total ARG abundance and richness^29^, yet selective retention or enrichment of ARGs, MGEs, and treatment-tolerant taxa has also been widely documented^30–32^. Because reductions in ARG abundance do not necessarily reflect equivalent reductions in dissemination potential or clinically relevant hazard, wastewater AMR studies that rely primarily on abundance- and composition-based analyses provide only limited insight into how treatment reshapes resistome risk.

Reflecting growing recognition of the environmental dimension of AMR, wastewater has become an increasingly prominent focus of international and national AMR policy frameworks. The World Health Organisation’s Global Action Plan on Antimicrobial Resistance identifies environmental pathways as an important component of a One Health response to AMR^33^, while the UK National Action Plan 2024-2029 highlights improved understanding of environmental transmission routes, wastewater surveillance, and evidence-based mitigation strategies as key research priorities^34^. Although the magnitude of environmentally mediated transmission remains uncertain, wastewater represents one of the few environmental compartments where large human-associated resistomes can be routinely monitored and where interventions capable of reducing ARG dissemination can be implemented at scale^35^. Consequently, improving methods that distinguish reductions in resistome burden from reductions in epidemiologically relevant resistome risk is essential for informing both surveillance and future wastewater management strategies.

Addressing this challenge requires analytical frameworks capable of integrating ARG identity with genomic context, host association, and mobility potential. MetaCompare2.0^36^ was developed to operationalise this concept by quantifying resistome risk as an emergent property of ARG-MGE-host configurations reconstructed from metagenomic assemblies. Rather than treating ARGs as isolated units, MetaCompare2.0 situates them within contig-level genomic contexts and incorporates evidence of MGE association and pathogen linkage to derive quantitative risk metrics. This framework distinguishes Ecological Resistome Risk (ERR), which reflects dissemination potential and ecological mobility within microbial communities, from Human Health Resistome Risk (HHRR), which reflects clinically relevant ARG-pathogen configurations associated with elevated public health concern. This therefore enables mechanistically informed comparison of resistome risk across environments.

Here, we apply MetaCompare2.0^36^ to a longitudinal collection of paired influent and effluent metagenomes obtained from nine UK WwTWs, sampled at six time-points within a year, to determine how wastewater treatment reshapes resistome risk profiles. We investigate how treatment influences the relative contributions of ecological dissemination risk and human health-associated risk across wastewater systems. This study advances beyond descriptive surveillance towards a more mechanistically informed understanding of wastewater treatment as an ecological filtering process within the environmental AMR landscape. More broadly, this work contributes to the development of scalable genomic frameworks that support comparative surveillance, environmental risk assessment, and evidence-based optimisation of wastewater treatment for AMR mitigation.

## Results

### Wastewater treatment restructures resistome diversity and composition

We first examined whether wastewater treatment altered ARG diversity and composition across 100 influent and final effluent metagenomes collected as 50 matched influent-effluent pairs. Shannon diversity was significantly lower in effluent than influent samples, based on matched influent-effluent comparisons (paired Wilcoxon signed-rank test, n = 50 pairs, p = 0.0176; Fig. 1A), with a moderate effect size (r = 0.34). Median Shannon diversity decreased from 4.22 in influent to 4.16 in effluent, corresponding to a median paired difference of −0.0783 Shannon units. Effluent samples showed greater variability in alpha diversity than influent samples (IQR: 0.31 vs. 0.11).

**Figure 1.**
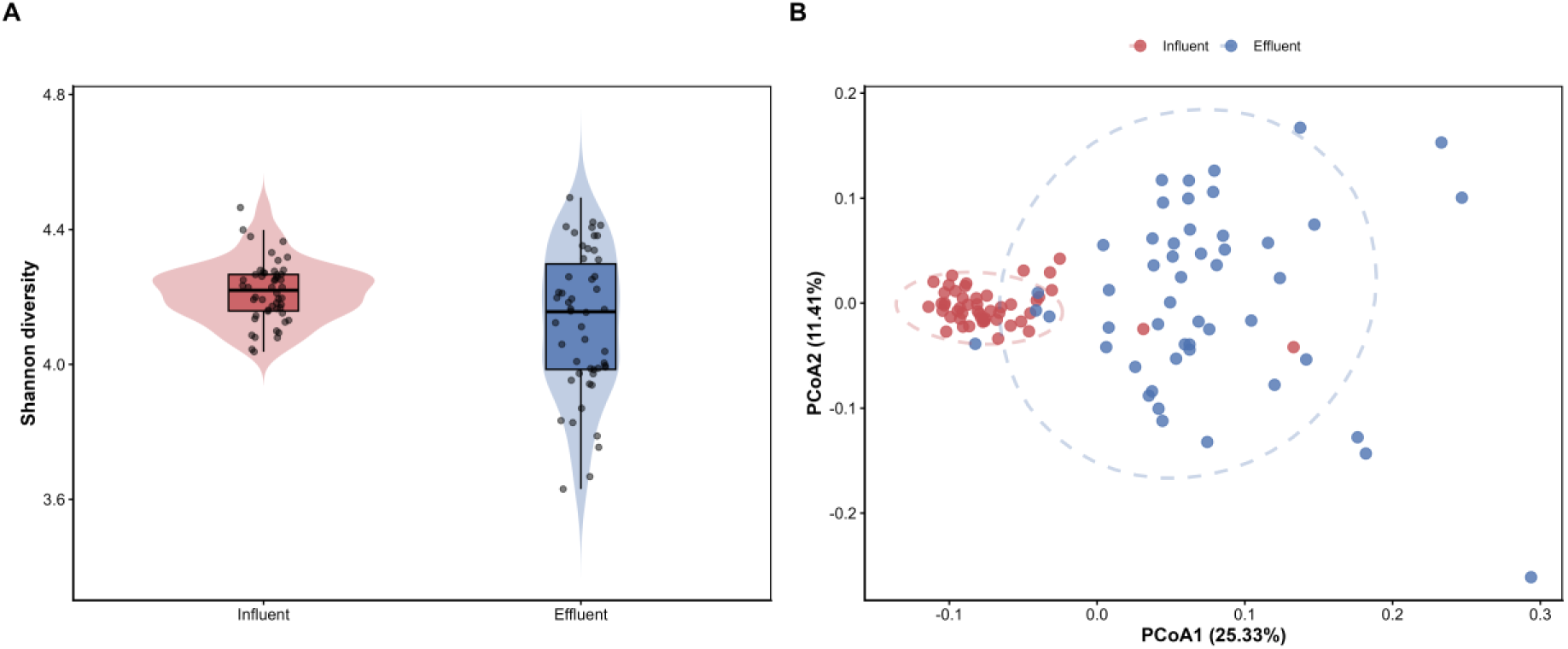
(A-B) ARG profiles were assessed across 100 metagenomes collected from nine wastewater treatment works (influent n = 50; effluent n = 50) at six timepoints within a year. (A) Violin and jitter plots display alpha diversity (Shannon index) of ARGs in influent and effluent samples. Points represent individual metagenomes, and embedded violins depict the distribution of Shannon indices. (B) Principal Coordinates Analysis (PCoA) of Bray-Curtis dissimilarities calculated from Hellinger-transformed relative ARG abundances illustrates beta diversity.

Wastewater treatment was associated with restructuring in resistome composition. Principal coordinates analysis (PCoA) based on Bray-Curtis dissimilarities revealed separation between influent and effluent resistomes (Fig. 1B). The first two axes explained 25.33% and 11.41% of total variation, respectively. Permutational multivariate analysis of variance (PERMANOVA), with permutations constrained by WwTW, identified treatment stage (i.e. influent/final effluent) as significantly associated with differences in resistome composition (R² = 0.166, *F* = 19.48, *P* = 0.001). Multivariate dispersion differed significantly between influent and effluent (betadisper followed by permutation testing, F = 68.98, permutation P = 0.001), indicating differences in within-group variability. These results indicate that wastewater treatment causes relatively modest changes in ARG alpha diversity but is associated with substantial restructuring of resistome composition, accompanied by increased between-sample variability among effluent communities. This pattern is consistent with ecological restructuring during treatment, where the resistome shifts substantially despite relatively small changes in overall ARG diversity.

### Wastewater treatment lowers ecological and human health resistome risk

We next evaluated whether treatment-associated shifts in resistome composition translate into changes in functional risk. We did this using MetaCompare2.0^36^. Two complementary metrics were considered: Ecological Resistome Risk (ERR), reflecting the co-localisation of ARGs with MGEs and thus their dissemination potential within microbial communities, and Human Health Resistome Risk (HHRR), reflecting ARG associations with clinically relevant host taxa. ERR shifted from broadly distributed values in influents to a tightly constrained low-risk state in effluents (Fig. 2A, D). A linear mixed-effects model with WwTW included as a random intercept confirmed a strong negative effect of treatment on ERR (β = −23.96, p < 1 × 10^-26^), consistent with reduced representation of ARG profiles associated with higher mobility potential. Paired analyses supported this pattern, with most matched influent-effluent pairs showing reduced ERR (median Δ = −25.35; Wilcoxon signed-rank test, p < 0.0001). A similar but attenuated pattern was observed for HHRR. Influent samples exhibited higher median values and greater dispersion, whereas effluent communities converged towards consistently low-risk states. Mixed-effects modelling again indicated a significant reduction associated with treatment (β = −1.69, p < 1 × 10^-23^), and paired comparisons confirmed a strong decrease across matched samples (median Δ = −1.70; p < 0.0001).

**Figure 2.**
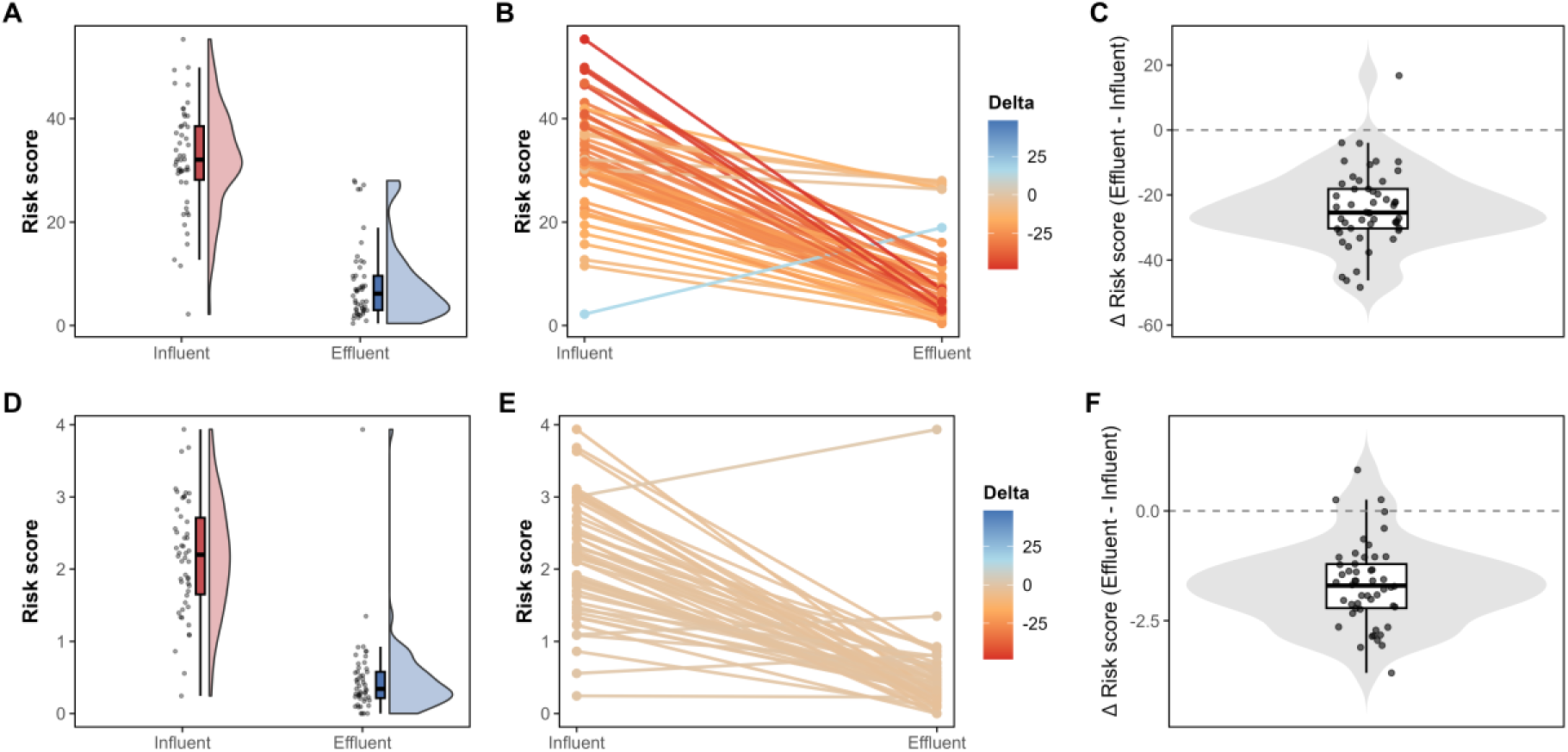
(A-C) Ecological Resistome Risk (ERR) and (D-F) Human Health Resistome Risk (HHRR) were quantified using MetaCompare2.0 across 100 metagenomes collected from nine wastewater treatment works (influent *n* = 50; effluent *n* = 50). (A, D) Raincloud plots display the distribution and density of ERR and HHRR scores in influent and effluent samples; points represent individual metagenomes, and embedded boxplots indicate median and interquartile range. (B, E) Paired influent-effluent comparisons for each sampling round and treatment plant, with each line representing a matched pair; line colour denotes the magnitude and direction of change. (C, F) Boxplots summarise paired differences (effluent-influent) in ERR and HHRR, with negative values indicating risk reduction following treatment.

Notably, reductions in both ERR and HHRR occurred despite the increased compositional dispersion observed in effluent resistomes, indicating that increased between-sample heterogeneity was not associated with increased functional risk. Instead, treatment appears to be associated with a consistent reduction in ARG profiles linked to mobility potential and clinically relevant hosts, while allowing greater variability among the remaining low risk resistome configurations.

### Wastewater treatment consistently reduces resistome risk despite site-specific variation

To determine whether treatment-associated reductions in resistome risk were consistent across individual WwTWs, we assessed ERR and HHRR changes within each site. Site-level paired comparisons showed negative shifts in both ERR and HHRR following treatment across all WwTWs. However, statistical significance at individual sites was limited by the small number of paired observations per WwTW (n = 5-6), and no site-level comparisons remained significant after Benjamini-Hochberg correction. To formally test whether the magnitude of risk reduction differed between the nine WwTWs, a two-way ANOVA (RiskScore ∼ Group × Site) was fitted for each risk metric (I.e., ERR/HHRR; Fig. 3). This revealed a significant Group × Site interaction for both ERR (F₈,₈₂ = 3.05, p = 0.0047) and HHRR (F₈,₈₂ = 2.55, p = 0.016), indicating that while all nine WwTWs showed a reduction in resistome risk following treatment, the magnitude of this reduction varied significantly among them. The treatment main effect nonetheless explained substantially more variance than the interaction (partial η² = 0.78 and 0.72 for ERR and HHRR, respectively, versus 0.23 and 0.20 for the Group × Site interaction), indicating that the overall treatment effect was considerably larger than site-to-site variation in its magnitude. Together, these results indicate that resistome risk attenuation was directionally consistent across the sampled wastewater treatment works but not uniform in magnitude.

**Figure 3.**
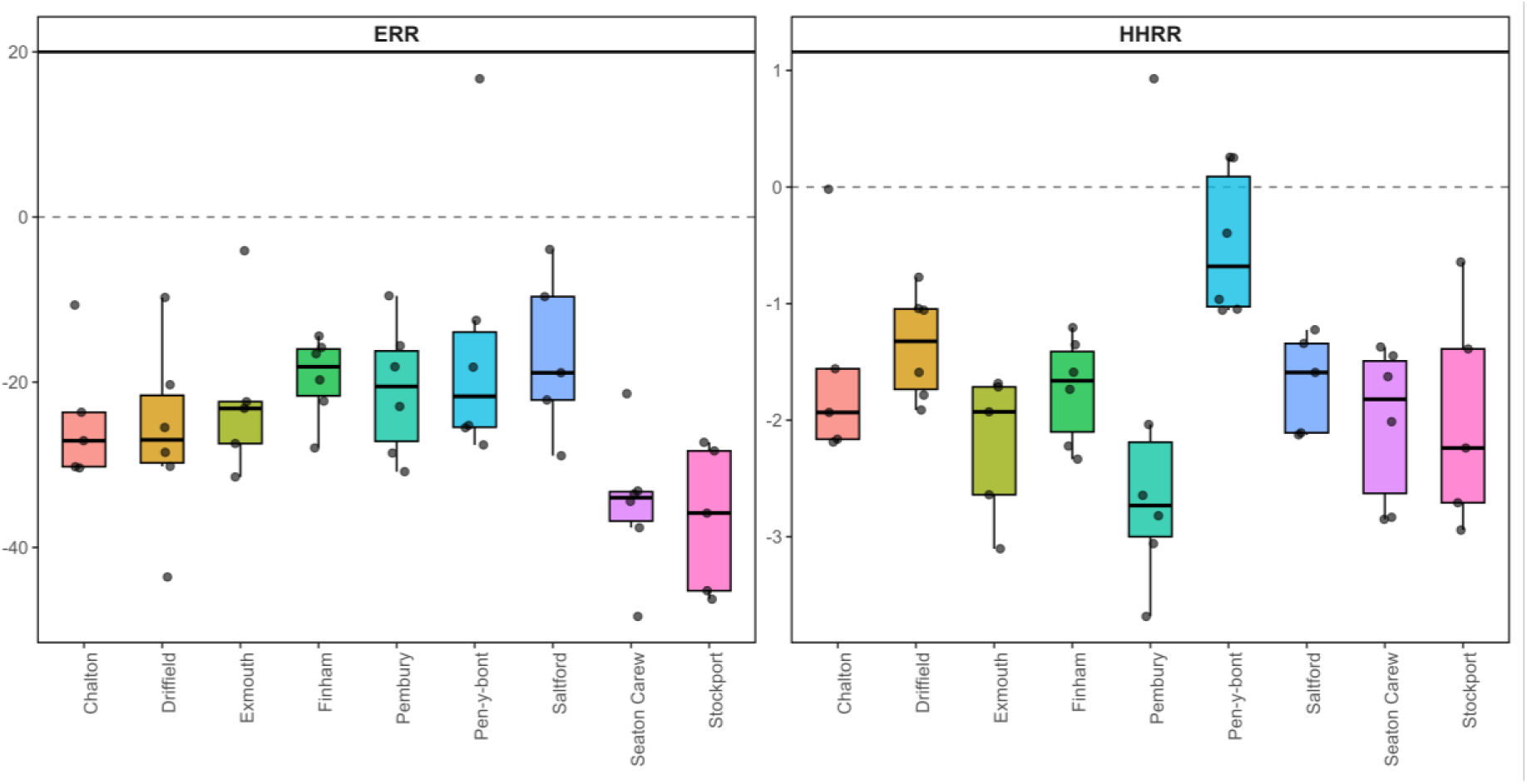
Paired change in Ecological Resistome Risk (ERR) and Human Health Resistome Risk (HHRR) following treatment (Influent-Effluent) at each of the nine wastewater treatment works. Boxplots show the distribution of paired plant-round differences per site; points represent individual matched pairs. A significant Group × Site interaction (two-way ANOVA) indicates that the magnitude of risk reduction differed significantly among sites for both ERR and HHRR.

Given this site-to-site variation in the magnitude of attenuation, we next assessed whether it was systematically associated with the secondary treatment technologies employed at each of these WwTWs, comparing activated sludge (AS), trickling filters (TF), biological aerated flooded filters (BAFF), and hybrid AS-TF systems. Linear mixed-effects models fitted separately for ERR and HHRR (RiskScore ∼ Group × Secondary, treatment site as a random intercept) showed a strong overall reduction in risk scores following treatment for both metrics (ERR: F₁,₈₇.₀ = 135.2, p = 2.08 × 10⁻¹⁹; HHRR: F₁,₈₆.₉ = 135.4, p = 2.05 × 10⁻¹⁹). There was no evidence that the magnitude of treatment-associated risk reduction differed among secondary treatment configurations for either metric (Group × Secondary interaction, ERR: F₃,₈₇.₀ = 1.24, p = 0.301; HHRR: F₃,₈₆.₉ = 1.49, p = 0.222). Secondary treatment configuration alone did not significantly explain variation in risk scores for either metric (ERR: F₃,₅.₂ = 1.41, p = 0.339; HHRR: F₃,₅.₂ = 1.04, p = 0.447), indicating that reductions in resistome risk were not strongly dependent on the specific secondary treatment process.

Consistent with this result, treatment-associated reductions in ERR and HHRR were comparable across secondary treatment configurations. Median ERR reductions ranged from −18.9 in trickling filter systems to −27.3 in activated sludge systems, while HHRR reductions ranged from −1.4 to −2.1 across configurations. Despite modest variation in the magnitude of reduction among treatment types, all configurations showed consistent decreases in both risk metrics. These findings indicate that attenuation of high-risk ARG configurations is likely a reproducible feature of wastewater treatment in general, rather than a property of any individual secondary treatment process.

To determine whether variation in resistome risk could be explained by bulk wastewater chemistry, we fitted mixed-effects models that related ERR and HHRR to individual physicochemical parameters while accounting for treatment stage and treatment site as a random intercept (a shared sampling-round random effect was also tested but was not statistically estimable given the number of distinct sampling rounds available). No associations remained significant following correction for multiple testing (all adjusted p > 0.05), and no measured physicochemical parameter significantly explained variation in ERR following correction. Treatment stage (i.e. influent/final effluent) remained the dominant predictor of both ERR and HHRR throughout the analysis.

### Assembly contiguity influences linkage recovery but does not account for treatment-associated risk attenuation

MetaCompare2.0 infers ARG-MGE-host linkage structures directly from assembled contigs; therefore, we evaluated whether systematic differences in assembly quality could bias risk estimates in this analysis across influent and final effluent samples. Assembly statistics were broadly comparable between influent and final effluent metagenomes. No significant differences were detected in N50 (Mann-Whitney U test, U = 1515.5, p = 0.0677), contig number (U = 1178.0, p = 0.6221), or total assembly length (U = 1339.0, p = 0.5418). Although N50 showed a weak tendency towards differences between treatment stages, there was no evidence of a systematic shift in assembly quality that could explain downstream differences in risk estimation.

To quantify the extent to which assembly contiguity influences MetaCompare2.0 outputs, we examined correlations between N50 and ERR/HHRR. ERR showed a positive association with N50 (Spearman’s ρ = 0.436, p = 5.9×10^-6^), indicating that more contiguous assemblies were associated with increased recovery of ARG-MGE-host linkage structures. HHRR also correlated positively with N50, though to a lesser degree (ρ = 0.352, p = 3.3×10^-4^), reflecting its narrower dynamic range and lower absolute values. These correlations confirm that assembly contiguity contributes to the sensitivity of linkage recovery for both ERR and HHRR estimation.

Assembly quality alone, however, did not explain the pronounced differences in risk between influent and effluent. When N50 (scaled to kilobases) and treatment stage were jointly included in linear models, treatment stage (i.e., influent/effluent) remained strongly associated with both risk metrics after accounting for assembly contiguity. For ERR, effluent samples exhibited substantially lower scores independent of assembly contiguity (β = −23.67, p < 0.001), and the model explained 72% of total variance (R² = 0.718). N50 exerted a significant positive effect (β = 3.51 per kb, p < 0.001), consistent with improved linkage recovery in more contiguous assemblies. For HHRR, N50 had a modest positive effect (β = 0.146 per kb, p = 0.012), yet treatment stage again accounted for most variation, with effluent samples showing markedly reduced HHRR relative to influent (β = −1.67, p < 0.001; R² = 0.626).

While assembly contiguity appears to modestly enhance the recovery of ARG-MGE-host linkage structures, it does not account for the strong and reproducible attenuation of resistome risk observed following wastewater treatment. Treatment stage was the dominant predictor of both ERR and HHRR scores in these sensitivity analyses, with substantially larger effect estimates than assembly contiguity, supporting the interpretation that observed reductions in resistome risk are unlikely to be explained by differences in assembly quality alone and are instead consistent with treatment-associated biological changes.

## Discussion

WwTWs are widely recognised as critical nodes in the environmental AMR continuum because they concentrate microbial biomass, ARGs, MGEs, pathogens, and diverse chemical stressors from human, industrial, and environmental sources^28^. The introduction of anthropogenic antimicrobials has accelerated the mobilisation of ARGs from natural reservoirs into clinically relevant bacteria and MGEs^3,4^, reinforcing the need for mitigation strategies that extend beyond clinical settings and explicitly address environmental reservoirs and ecological processes^5^. Although metagenomic surveillance has advanced the characterisation of wastewater resistomes and enabled detailed descriptions of ARG diversity, abundance, and taxonomic distribution ^22,37^, the field still relies heavily on abundance-based endpoints that fail to capture dissemination potential or clinical hazard, risking mischaracterisation of epidemiological risk when mobile or clinically relevant ARG configurations persist^14,29^. This reflects a broader challenge in environmental AMR surveillance, where exposure to ARGs or resistant bacteria rarely produces immediate health outcomes: rather, it contributes to risk only after ecological transmission, HGT, colonisation, and eventual treatment failure, with ARGs persisting across long timescales and transitioning among hosts before reaching human or animal pathogens^18,19^. As a result, the epidemiological significance of an ARG is context-dependent and shaped by genomic context, host association, mobility, persistence, exposure opportunity, and onward transmission dynamics^21,22,25^. The lack of standardised methodologies that integrate these dimensions remains a major barrier to embedding genomics within routine AMR surveillance frameworks^38,39^.

Here, using a longitudinal, multisite dataset spanning nine WwTWs and one hundred paired influent-effluent metagenomes, we applied MetaCompare2.0 to distinguish ecological dissemination potential from pathogen-associated hazard by evaluating ARGs not only by their abundance but by their genomic configuration, that is, whether they are embedded within MGEs, co-located with other resistance determinants, or associated with pathogenic hosts, through quantification of ERR and HHRR^36^. The scale and replication of our sampling, spanning nine WwTWs, support an inference that is robust to site-level heterogeneity. A shared sampling-round random effect was also tested in all mixed-effects models but was not separately estimable given the number of distinct sampling occasions available, so temporal variability across rounds is addressed here through the paired-sampling design and paired Wilcoxon sensitivity analyses rather than through a dedicated round-level variance component^40^.

Across sites, influent ERR was consistently higher and substantially more dispersed than HHRR, indicating that incoming AMR pressure is dominated by heterogeneous reservoirs of potentially mobile ARGs, the magnitude of which likely reflects variable upstream anthropogenic inputs and microbial community composition^41^. In contrast, ARG compositions associated with clinically relevant pathogen signatures constituted a lower-magnitude, less variable subset of the broader resistome. These findings are consistent with human faecal inputs acting as concentrated reservoirs of pathogen-associated resistance determinants that subsequently undergo dilution and ecological turnover during conveyance through sewer networks^42,43^.

The observed separation between ERR and HHRR further demonstrates that ecological dissemination potential and clinically contextualised hazard are not interchangeable properties of the resistome but rather represent distinct, only partially overlapping, dimensions of AMR risk. This distinction is important because environmental AMR surveillance frequently treats all ARGs as epidemiologically equivalent despite substantial differences in their mobility, host range, and likelihood of contributing to clinically consequential transmission events^21,24^. Importantly, although ERR and HHRR are reported as risk metrics, they do not directly quantify adverse health outcomes. Rather, they are best interpreted as indicators of relative epidemiological relevance, since progression from environmental ARG occurrence to realised health risk depends on numerous intervening ecological and epidemiological processes. Moreover, HHRR occupies a narrower dynamic range than ERR and yields consistently lower absolute values; the comparatively modest HHRR reductions observed here should therefore be interpreted as relative shifts within this constrained scale rather than as direct comparisons in magnitude with ERR.

Both risk metrics declined across nearly all influent-effluent pairs, with consistent attenuation observed across all nine WwTWs. Because ERR and HHRR explicitly capture ARG-MGE-host configurations, these reductions are more consistent with treatment-driven restructuring of higher-risk genetic contexts than with simple dilution of ARG abundance. This distinction is particularly important because ARGs are not analogous to conventional chemical pollutants whose risk generally scales predictably with concentration^16^. Rather, AMR dissemination depends on dynamic ecological interactions involving host compatibility, selection pressure, HGT, and microbial community structure. Consequently, reductions in ARG abundance alone may not correspond proportionally to reductions in dissemination potential or clinically relevant hazard.

An important finding was that attenuation of resistome risk was consistent across AS, TF, BAFF, and hybrid AS-TF systems. Although treatment technologies differ substantially in engineering design and microbial ecology, no evidence was found that any configuration consistently outperformed the others with respect to ERR or HHRR reduction. This suggests that attenuation of high-risk ARG configurations may represent a general property of biological wastewater treatment rather than a feature of a particular secondary treatment process.

Notably, wastewater treatment substantially reduced ERR variability, suggesting preferential disruption of highly mobile ARG reservoirs that disproportionately contribute to influent heterogeneity. Residual ecological dissemination potential nevertheless remained detectable in effluent, indicating incomplete elimination of mobile ARG configurations. In contrast, HHRR declined more modestly, suggesting that pathogen-associated ARG carriage may be comparatively more persistent under treatment-associated selective pressures. This asymmetry may reflect the survival of specific taxa, protection within flocs or biofilms, the persistence of low-abundance resistant populations, or the chromosomal integration of resistance determinants that reduces dependence on MGEs for maintenance^14,30,44^.

Compositional and configuration-based endpoints exhibited markedly divergent behaviour across treatment. While influent resistomes clustered relatively tightly in ordination space, effluent resistomes became increasingly dispersed, indicating substantial site-specific reorganisation following treatment. Despite this increased compositional heterogeneity, both ERR and HHRR converged towards consistently lower and less variable values in effluent. One plausible interpretation is that WwTWs function as ecological filtering systems that selectively constrain dissemination-compatible resistance configurations while permitting persistence of broader background resistome diversity. Greater microbial diversity and niche competition within downstream aquatic communities have previously been proposed to suppress invasion and horizontal transfer of resistant organisms through mechanisms of niche exclusion and ecological resistance^45,46^. Under this framework, the persistence of ARGs following treatment does not necessarily imply equivalent persistence of mobilisable or clinically consequential resistance configurations. Accordingly, treated effluent should not be viewed as equivalent to untreated wastewater inputs. Although residual ERR and HHRR remained detectable after treatment, effluent represented a substantially attenuated and less variable AMR signal. Consequently, untreated wastewater is likely to introduce higher and more heterogeneous dissemination-compatible ARG configurations into receiving waters, whereas treated effluent represents a reduced, but not eliminated, source of AMR-relevant genetic contexts.

These findings help reconcile previous reports describing persistence of ARGs following wastewater treatment^14,32,47^ with observations of reduced downstream AMR risk. Specifically, our results demonstrate that compositional persistence does not necessarily equate to persistence of high-risk genetic contexts. This distinction is critical because commonly used ARG risk-ranking approaches often classify ARGs based on historical worst-case associations with pathogens and MGEs, irrespective of their actual genomic context within a given environmental sample^48^. Such approaches may overestimate epidemiological risk where ARGs are chromosomally linked to non-pathogenic environmental hosts that lack plausible transmission pathways to humans or animals^21^. Configuration-aware frameworks such as MetaCompare2.0 therefore provide a more mechanistic and epidemiologically grounded basis for interpreting environmental AMR surveillance data than abundance or composition alone.

Several methodological limitations inherent to short-read metagenomics should be considered when interpreting configuration-based resistome risk. Recovery of ARG genomic context from environmental metagenomes remains technically challenging for several reasons. For example, uneven species abundance distributions, repeated mobile elements, and the occurrence of ARGs across multiple genomic backgrounds^49,50^. ARG-containing contigs are particularly prone to assembly fragmentation because conserved ARG regions frequently occur across multiple genomic contexts, leading to breaks that disrupt inferred ARG-MGE-host linkage^51^. Environmental metagenomes are especially difficult to assemble because they contain highly diverse microbial communities comprising numerous closely related strains and low-abundance taxa^52,53^. Moreover, strain-level differentiation of pathogenic and non-pathogenic variants within disease-associated taxa typically requires whole-genome resolution or long-read sequencing, which is beyond the scope of this study. Sequencing depth also constrains sensitivity, particularly for low-abundance ARGs and MGEs that may remain undetected even in deeply sequenced datasets^54,55^. Although approximately 20 million reads per sample were analysed here, previous work suggests that substantially greater sequencing depth may still fail to fully capture ARG diversity in wastewater effluent^54^. In addition, methodological decisions regarding read mapping, assembly strategy, and ARG counting approaches can substantially influence the inferred resistome structure and functional interpretation^56^. Inferred risk configurations should therefore be interpreted as probabilistic representations of dissemination potential rather than definitive reconstructions of transmission pathways. Nevertheless, ongoing advances in assembly algorithms, integration of long-read sequencing, and context-aware bioinformatic frameworks continue to improve the biological interpretability of environmental resistome surveillance.

Consistent with these methodological considerations, we observed that MetaCompare2.0 risk estimates were partially influenced by assembly contiguity, with higher N50 values associated with increased recovery of ARG-MGE-host configurations. This pattern was evident for both ERR and HHRR metrics, although the strength of association differed. ERR showed a moderately significant correlation with N50. HHRR exhibited a weaker but still significant relationship. These trends are expected, as improved assembly continuity increases the likelihood of reconstructing multielement genomic contexts required for configuration-aware inference^57^. Crucially, however, assembly metrics did not differ systematically between influent and effluent samples, and treatment-associated reductions in both ERR and HHRR remained highly significant after adjusting for N50 effects. Thus, while assembly quality contributes to quantitative variation in inferred risk scores, it does not account for the consistent and substantial attenuation of resistome risk observed across treatment systems. Wastewater treatment appears to reliably reduce both ERR and HHRR, independent of variation in assembly contiguity. For the same reason, absolute ERR and HHRR values are conditional on assembly performance and the specific bioinformatic pipeline applied; they are therefore most appropriately compared within a consistently processed dataset, such as ours, rather than across studies using different sequencing depths, assemblers, or reference databases.

A major strength of this study is the reproducibility of risk attenuation across multiple WwTWs and repeated longitudinal sampling intervals. This consistency supports the interpretation that conventional wastewater treatment processes reproducibly reduce resistome risk rather than producing isolated or secondary treatment-type-specific effects. The discrepancies in reductions between specific WwTWs is possibly attributable to differing population equivalents, and primary/tertiary treatment types of the WwTWs not examined in this study, warranting further investigation.

From an operational perspective, distinguishing ERR from HHRR provides a more precise framework for evaluating treatment performance because different treatment processes may affect ARG mobility, microbial survival, and pathogen-associated carriage in different ways. This distinction has important practical implications, as optimisation strategies focused solely on reducing bulk ARG abundance may fail to target the specific ARG-MGE-host configurations most relevant to downstream epidemiological risk.

Overall, our findings support a conceptual shift in how wastewater systems are interpreted within the environmental AMR continuum. Rather than functioning solely as passive conduits or reservoirs of resistance pollution, WwTWs appear to operate as ecological selection and restructuring systems that differentially modulate dissemination-compatible ARG configurations. While resistance determinants persist beyond treatment, the consistent attenuation of ARG-MGE-host configurations associated with elevated dissemination potential and pathogen linkage indicates that treatment substantially reshapes the epidemiological relevance of the resistome without necessarily eliminating ARGs outright. More broadly, these findings support a transition from descriptive resistome surveillance towards mechanistic, configuration-aware evaluation of environmental AMR risk. Future work should integrate genomic risk metrics with process-level operational parameters, exposure assessment frameworks, and quantitative microbial risk assessment approaches to determine which treatment features most effectively disrupt ARG mobility and pathogen-associated resistance configurations, thereby enabling evidence-based optimisation of wastewater treatment for environmental AMR mitigation.

## Methods

### Study design and metadata collection

Wastewater samples were collected from nine WwTWs across England and Wales between September 2020 and July 2021 under the UK Chemical Investigation programme (CIP), operated by 10 water and wastewater companies in England and Wales, in collaboration with regulators (Defra, the Environment Agency and Natural Resources Wales). Wastewater treatment works (WwTWs) were selected to represent a range of catchment sizes, population equivalents, and treatment configurations, including activated sludge, trickling filter, and tertiary polishing processes. Each site was sampled six times over an 11-month period, generating paired raw influent and final effluent samples per sampling round (total n = 100 metagenomes; 50 influent, 50 effluent).

Operational metadata were collated for each sampling event and included dry weather flow, full flow to treatment, hydraulic retention time, sludge retention time, treatment configuration, and chemical dosing regimens where available. Physicochemical measurements included pH, biochemical oxygen demand (BOD), chemical oxygen demand (COD), ammoniacal nitrogen, nitrate, nitrite, chloride, dissolved organic carbon, total phosphorus, total dissolved phosphorus, and suspended solids, using methods described in Bowes et al., 2018^58^. Metadata were curated and harmonised prior to analysis.

### Sample collection and preservation

Liquid samples (raw influent and final effluent) were collected using 24-hour time-proportional composite samplers programmed to collect aliquots at 30-minute intervals. Approximately 10 L per composite was obtained per 24-h sampling event. Samples were homogenised immediately after collection.

Subsamples for molecular analysis were transferred into sterile high-density polyethylene bottles and transported on dry ice to the laboratory within 24 h. Samples were stored at −80 °C until processing. Field blanks and extraction blanks were included to monitor contamination.

### Biomass concentration and DNA extraction

Microbial biomass was concentrated by filtering 250-500 mL of wastewater through a sterile 0.22 μm pore-size MF-Millipore membrane filter (Merck Millipore Darmstadt, Germany; Cat. GSWP04700). Filtering equipment was thoroughly cleaned between each sample. Following filtration, the membrane was removed with ethanol sterilised tweezers, cut into pieces with ethanol sterilised scissors, and transferred into ZR BashingBead™ Lysis Tubes (Zymo Research, Irvine, CA, USA, Cat. S6003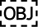50) containing 750 µL DNA/RNA Shield (Zymo Research, Irvine, CA, USA; Cat. R1100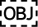250). The total volume of liquid filtered was recorded for subsequent analyses. Samples in the BashingBead™ tubes were mechanically lysed using a FastPrep instrument (QBioGene, Irvine, CA, USA) at 6,000 rpm for 5 minutes and then stored at −20 °C until DNA extraction.

DNA was extracted using the Zymo Research Quick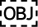DNA Faecal/Soil Microbe Kit (Zymo Research, Irvine, CA, USA, Cat. D6010). Extractions were carried out according to the manufacturer’s protocol, with the following modifications: extraction commenced from step 3 of the protocol (steps 1 and 2 had been completed before freezing), and DNA/RNA Shield was used in place of the BashingBead™ Buffer, in line with the manufacturer’s recommended alternative. The centrifugation step at stage 9 was extended by an additional 30 seconds to ensure removal of residual ethanol.

DNA was eluted in 100 µL of the kit’s elution buffer. DNA yield and quality were assessed using a NanoDrop™ 8000 Spectrophotometer (NanoDrop Technologies, Wilmington, DE, USA). DNA concentration was quantified using the Qubit® dsDNA BR Assay Kit in combination with a Qubit® 3.0 Fluorometer (Invitrogen, Carlsbad, CA, USA). All DNA extracts were stored at −20 °C until further analysis.

### Shotgun metagenomic library preparation and sequencing

Metagenomic libraries were prepared using the NEBNext Ultra II DNA Library Prep Kit for Illumina (New England Biolabs). Briefly, 100 ng of input DNA was enzymatically fragmented to a target insert size of ∼350 bp, followed by end repair, A-tailing, and ligation of dual-index adapters. Libraries were purified using AMPure XP beads (Beckman Coulter) and amplified by 6-8 cycles of PCR. Library fragment size distributions were assessed using an Agilent 2100 Bioanalyzer (High Sensitivity DNA kit). Libraries were quantified using qPCR (KAPA Library Quantification Kit, Roche). Equimolar library pools were sequenced on an Illumina NovaSeq 6000 platform (2 × 150 bp paired-end reads), generating a median sequencing depth of approximately 6 Gbp per sample at Novogene. Negative controls were sequenced and screened to confirm absence of significant contaminant signal.

### Sequence quality control and assembly

Raw reads were processed using fastp^59^ v0.23.2 with parameters --detect_adapter_for_pe, --cut_front, --cut_tail, --cut_mean_quality 30, and minimum read length of 50 bp. Quality filtering reports were generated for each sample. Host contamination was screened using Bowtie2^60^ v2.5.1 in end-to-end mode against the human reference genome (GRCh38), and mapped reads were removed. Quality-controlled reads were assembled de novo using MEGAHIT^61^ v1.2.9 with default meta-sensitive parameters (--k-min 21 --k-max 141 --k-step 12). Contigs shorter than 500 bp were discarded. Assembly statistics were generated using QUAST^62^ v5.2.0. Read mapping back to assemblies was performed using Bowtie2 v2.5.1, and coverage statistics were calculated with Samtools^63^ v1.17.

### Resistome risk profiling

ERR and HHRR were quantified using MetaCompare2.0 v1^33^. The pipeline was installed from the official GitHub repository (<u>GitHub - mrumi/MetaCompare2.0 ·GitHub</u>), and the accompanying metacmpDB reference database (Zenodo record 10626079) was used without modification.

MetaCompare2.0 identifies ARGs, MGEs, and pathogen-associated sequences within assembled contigs. ARGs are detected using sequence similarity searches against curated resistance databases with default thresholds (≥80% identity and ≥70% coverage). MGEs are identified based on integrase, transposase, and plasmid-associated markers, while pathogen associations are assigned via alignment to reference genomes in metacmpDB.

For each sample, contigs are classified into four categories based on the presence of annotated features: (i) contigs containing one or more ARGs, (ii) contigs containing one or more ARGs and one or more MGEs, (iii) contigs containing one or more ARGs, MGEs, and pathogen-associated sequences, and (iv) contigs containing one or more ARGs and pathogen-associated sequences. These categories are not mutually exclusive. The proportions of contigs in each category are calculated by normalising counts to the total number of contigs (N*_contigs_*), yielding four metrics: Q_ARG_, Q_ARG,MGE_, Q_ARG,MGE,PAT_, and Q_ARG,PAT_.

ERR is computed in a three-dimensional hazard space defined by Q_ARG_, Q_ARG,MGE_, and Q_ARG,MGE,PAT_, while HHRR incorporates an additional fourth dimension, Q_ARG,PAT_. ERR considers the full set of detected ARGs and pathogen annotations, whereas HHRR is restricted to clinically relevant Rank I ARGs^48^ and ESKAPEE-associated pathogens as defined in metacmpDB.

The Euclidean distance of each sample to the theoretical maximum point (all dimensions equal to 1) is calculated as:

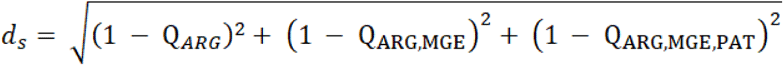

(for ERR), and:

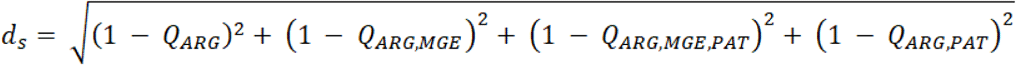

(for HHRR).

The normalisation factor (d_W_), corresponding to a sample with no ARGs (all Q = 0), is given by: d_w_ = √3 (for ERR), and:

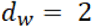

(for HHRR).

The final risk score is calculated as:

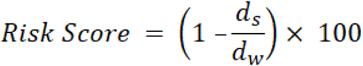

This formulation constrains scores between 0 and 100, where higher values indicate greater resistome risk due to increased ARG abundance, mobility potential, and association with human pathogens ^36^.

All samples were processed using default scoring parameters. Workflow execution was managed using Snakemake v7.32.4, with Conda-managed environments ensuring reproducibility.

### ARG data processing and diversity analyses

ARG count data were imported in long format and converted to a sample-by-feature abundance matrix. Samples with zero total ARG counts were removed, and counts were converted to relative abundances using total-sum scaling (decostand(method = “total”), vegan^64^ v2.6-4).

Shannon diversity (diversity()) was calculated on the full relative-abundance dataset, prior to prevalence filtering, and averaged within treatment plant × sampling round × treatment stage combinations. Matched influent-effluent comparisons used paired Wilcoxon signed-rank tests.

For beta-diversity analyses, ARGs detected in fewer than 5% of samples were excluded. The resulting dataset was Hellinger-transformed before calculating Bray-Curtis dissimilarities (vegdist()), and PCoA was performed using cmdscale(). Differences in composition between influent and effluent were assessed using PERMANOVA (adonis2(), 999 permutations), with permutations restricted within WwTW. Homogeneity of multivariate dispersion was assessed using betadisper() followed by permutest(), with the same restricted permutation structure.

### Statistical analysis and visualisation

All analyses were performed in R 4.3.2. Mixed-effects models were fitted using lme4 and lmerTest, diagnostics using performance, and figures using ggplot2 with cowplot, patchwork, ggpubr, ggrain and ggtext (600 dpi).

All mixed-effects models were initially fitted with crossed random intercepts for treatment plant/site, sampling round, and their interaction. Where this structure resulted in singular fits, models were refitted using alternative optimisers (bobyqa, Nelder-Mead) before simplifying the random-effects structure. Simplified models retained treatment plant/site as a random intercept. Mixed-effects models served as the primary inferential analyses throughout. Paired Wilcoxon signed-rank tests, following averaging of replicates within each plant/site × round × stage combination, served as secondary, distribution-free sensitivity analyses confirming the direction of effects. P-values were Benjamini-Hochberg adjusted within each family of comparisons.

MetaCompare2.0 ERR and HHRR scores were modelled with treatment stage as a fixed effect (RiskScore ∼ Group + (1|Plant) + (1|Round) + (1|Plant:Round)); as above, this produced singular fits for both metrics and was simplified to a treatment-plant random intercept alone.

Consistency across individual WwTWs was examined using paired Wilcoxon tests per site (same averaging as above), and a two-way ANOVA (RiskScore ∼ Group × Site, both fixed), fitted separately per metric, tested whether the magnitude of reduction differed between WwTWs; partial η² was calculated from the model sums of squares to compare the relative contribution of the treatment effect and the interaction.

To assess variation by secondary treatment technology, WwTWs were classified into four categories (activated sludge, AS; trickling filters, TF; biological aerated flooded filters, BAFF; hybrid AS-TF), with treatment stage, secondary treatment configuration, and their interaction as fixed effects (RiskScore ∼ Group × Secondary); as above, this produced singular fits for both metrics and was simplified to a treatment-site random intercept. Estimated marginal means (emmeans) and treatment contrasts across configurations, pooled across both metrics, were Benjamini-Hochberg adjusted.

Associations between physicochemical parameters and risk scores were assessed using linear mixed-effects models fitted separately per variable, with the variable and, where appropriate, treatment stage as fixed effects; as above, this produced singular fits for every variable and was simplified to a treatment-site random intercept throughout. Values below the detection limit were replaced with one-half the detection limit; selected variables were log10-transformed, pH was untransformed. Pairwise Spearman correlations among predictors assessed redundancy, since each was modelled independently.

### Assessment of assembly quality effects on MetaCompare risk scores

Assembly quality (N50, contig number, total assembly length) was compared between influent and effluent samples using two-sided Mann-Whitney U tests, and its association with risk scores assessed using Spearman correlations between N50 and ERR/HHRR separately. Exploratory OLS models (RiskScore ∼ Group + N50_kb, N50 scaled to kilobases), fitted separately for ERR and HHRR, tested as sensitivity analyses whether assembly quality explained treatment-associated differences in risk. All tests were two-sided.

## Author Contributions

RE: Data curation, formal analysis, investigation, visualisation, writing-original draft preparation; SBB: Conceptualisation, data curation, formal analysis, investigation, visualisation, resources, supervision, validation, writing-original draft preparation; LKN: Investigation, resources, writing-original draft preparation, Writing – Review & Editing; MJB: Investigation, resources, writing-original draft preparation, Writing – Review & Editing; LKA: Resources, writing-original draft preparation; DJEN: Resources, writing-original draft preparation; HSG: data curation, investigation, resources, validation, writing-original draft preparation; BKH: data curation, formal analysis, investigation, resources, validation, funding acquisition, writing-original draft preparation; DSR: Conceptualisation, data curation, formal analysis, investigation, visualisation, resources, supervision, validation, funding acquisition, project administration, writing-original draft preparation; HT: Conceptualisation, data curation, formal analysis, investigation, visualisation, resources, supervision, validation, funding acquisition, project administration, writing-original draft preparation.

## Data Availability

The source code and data files for this analysis can be found at https://github.com/UKCEH-MolecularEcology/arg-rank-cip3.

