## Supplementary materials for "Wastewater Treatment Attenuates Ecological and Human Health Resistome Risk"

S1. Alpha diversity summary

| Stage | n | median | IQR | mean | sd |
|---|---|---|---|---|---|
| Influent | 50 | 4.219687 | 0.108147 | 4.215237 | 0.090849 |
| Effluent | 50 | 4.156072 | 0.31283 | 4.122229 | 0.211907 |

S2. Wilcoxon results

| n_paired | median_influent | median_effluent | median_difference | wilcoxon_p | ci_lower | ci_upper | effect_size_r |
|---|---|---|---|---|---|---|---|
| 50 | 4.219687 | 4.156072 | -0.07832 | 0.017563 | -0.15759 | -0.01436 | 0.336517 |

S3. PCoA Variance explained

| Axis | Variance_Explained_Percent |
|---|---|
| PCoA1 | 25.33 |
| PCoA2 | 11.41 |
| PCoA3 | 9.19 |
| PCoA4 | 6.6 |
| PCoA5 | 4.38 |

S4. PERMANOVA

| Df | SumOfSqs | R2 | F | Pr(>F) | Term |
|---|---|---|---|---|---|
| 1 | 0.545819 | 0.16582 | 19.4807 | 0.001 | Model |
| 98 | 2.745809 | 0.83418 | NA | NA | Residual |

| | | | | | |
|---|---|---|---|---|---|
| 99 | 3.291628 | 1 | NA | NA | Total |

S5. Betadisperser permutation

| Df | Sum Sq | Mean Sq | F | N.Perm | Pr(>F) | Term |
|---|---|---|---|---|---|---|
| 1 | 0.218008 | 0.218008 | 68.97694 | 999 | 0.001 | Groups |
| 98 | 0.309738 | 0.003161 | NA | NA | NA | Residuals |

S6. MC2 Mixed Model

| effect | term | estimate | std.error | statistic | df | p.value | conf.low | conf.high | ScoreTypeLabel | R2_marginal | R2_conditional | p.adj |
|---|---|---|---|---|---|---|---|---|---|---|---|---|
| fixed | GroupEffluent | -23.9566 | 1.536612 | -15.5905 | 90.15115 | 2.50E-27 | -27.0092 | -20.9039 | ERR | 0.634033 | 0.741758 | 5.00E-27 |

| | | | | | | | | | | | | |
|---|---|---|---|---|---|---|---|---|---|---|---|---|
| fixed | GroupEffluent | -1.68573 | 0.12275 | -13.733 | 90.24586 | 7.92E-24 | -1.92958 | -1.44187 | HHRR | 0.595389 | 0.68746 | 7.92E-24 |

S7. MC2 Paired wilcoxon

| ScoreTypeLabel | n_pairs | Median_Influent | Median_Effluent | Median_Delta | Wilcoxon_p | CI_lower | CI_upper | effect_size_r | p.adj |
|---|---|---|---|---|---|---|---|---|---|
| ERR | 50 | 32.05416 | 6.164326 | -25.3537 | 1.61E-09 | -27.4005 | -21.3232 | 0.85392 | 1.61E-09 |
| HHRR | 50 | 2.199954 | 0.341333 | -1.70072 | 1.61E-09 | -1.97067 | -1.47065 | 0.85392 | 1.61E-09 |

S8. Site-level paired wilcoxon

| Site | ScoreType | n_pairs | p | p.adj |
|---|---|---|---|---|
| Chalton | ERR | 5 | 0.0625 | 0.070313 |

| | | | | |
|---|---|---|---|---|
| Chalton | HHRR | 5 | 0.0625 | 0.070313 |
| Driffield | ERR | 6 | 0.03125 | 0.070313 |
| Driffield | HHRR | 6 | 0.03125 | 0.070313 |
| Exmouth | ERR | 5 | 0.0625 | 0.070313 |
| Exmouth | HHRR | 5 | 0.0625 | 0.070313 |
| Finham | ERR | 6 | 0.03125 | 0.070313 |
| Finham | HHRR | 6 | 0.03125 | 0.070313 |
| Pembury | ERR | 6 | 0.03125 | 0.070313 |
| Pembury | HHRR | 6 | 0.0625 | 0.070313 |
| Pen-y-bont | ERR | 6 | 0.09375 | 0.09375 |
| Pen-y-bont | HHRR | 6 | 0.15625 | 0.15625 |
| Saltford | ERR | 5 | 0.0625 | 0.070313 |

| | | | | |
|---|---|---|---|---|
| Saltford | HHRR | 5 | 0.0625 | 0.070313 |
| Seaton Carew | ERR | 6 | 0.03125 | 0.070313 |
| Seaton Carew | HHRR | 6 | 0.03125 | 0.070313 |
| Stockport | ERR | 5 | 0.0625 | 0.070313 |
| Stockport | HHRR | 5 | 0.0625 | 0.070313 |

S9. Site interaction ANOVA

| Df | Sum Sq | Mean Sq | F value | Pr(>F) | Term | ScoreType |
|---|---|---|---|---|---|---|
| 1 | 14347.91 | 14347.91 | 287.0874 | 1.63E-28 | Group | ERR |
| 8 | 2749.407 | 343.6759 | 6.876611 | 6.91E-07 | Site | ERR |
| 8 | 1218.991 | 152.3739 | 3.048849 | 0.004717 | Group:Site | ERR |

| | | | | | | |
|---|---|---|---|---|---|---|
| 82 | 4098.156 | 49.97751 | NA | NA | Residuals | ERR |
| 1 | 71.04192 | 71.04192 | 214.2158 | 1.39E-24 | Group | HHRR |
| 8 | 13.31968 | 1.66496 | 5.020426 | 4.36E-05 | Site | HHRR |
| 8 | 6.755071 | 0.844384 | 2.546107 | 0.015742 | Group:Site | HHRR |
| 82 | 27.19425 | 0.331637 | NA | NA | Residuals | HHRR |

S10. Treatment type ANOVA

| Sum Sq | Mean Sq | NumDF | DenDF | F value | Pr(>F) | Term | ScoreType |
|---|---|---|---|---|---|---|---|
| 7924.456 | 7924.456 | 1 | 87.03343 | 135.2223 | 2.07538044392193e-19 | Group | ERR |
| 248.3992 | 82.79974 | 3 | 5.221175 | 1.412889 | 0.338793 | Secondary | ERR |

| | | | | | | | |
|---|---|---|---|---|---|---|---|
| 217.7087 | 72.56955 | 3 | 87.03343 | 1.238322 | 0.300768 | Group:Secondary | ERR |
| 50.24542 | 50.24542 | 1 | 86.94604 | 135.3565 | 2.05117058186088e-19 | Group | HHRR |
| 1.163223 | 0.387741 | 3 | 5.161734 | 1.044538 | 0.447162 | Secondary | HHRR |
| 1.664206 | 0.554735 | 3 | 86.94604 | 1.494405 | 0.221722 | Group:Secondary | HHRR |

S11. Treatment type contrasts

| contrast | Secondary | estimate | SE | df | t.ratio | p.value | ScoreType | p.adj |
|---|---|---|---|---|---|---|---|---|
| Effluent - Influent | AS | -26.1478 | 2.010372 | 87.00079 | -13.0065 | 4.14E-22 | ERR | 3.31E-21 |
| Effluent - Influent | TF | -19.0035 | 3.264218 | 87.00079 | -5.82176 | 9.55E-08 | ERR | 1.91E-07 |

| | | | | | | | | |
|---|---|---|---|---|---|---|---|---|
| Effluent - Influent | BAFF | -21.705 | 4.841618 | 87.00079 | -4.48302 | 2.23E-05 | ERR | 2.55E-05 |
| Effluent - Influent | AS_TF | -24.3954 | 4.841618 | 87.00079 | -5.03869 | 2.52E-06 | ERR | 3.36E-06 |
| Effluent - Influent | AS | -1.50464 | 0.160002 | 87.00107 | -9.40393 | 6.69E-15 | HHRR | 2.67E-14 |
| Effluent - Influent | TF | -1.97402 | 0.259793 | 87.00107 | -7.59842 | 3.22E-11 | HHRR | 8.58E-11 |
| Effluent - Influent | BAFF | -2.21477 | 0.385335 | 87.00107 | -5.74764 | 1.31E-07 | HHRR | 2.10E-07 |
| Effluent - Influent | AS_TF | -1.57273 | 0.385335 | 87.00107 | -4.08146 | 9.90E-05 | HHRR | 9.90E-05 |

S12. Treatment type paired summary

| ScoreType | Secondary | n | median_delta | IQR_delta | mean_delta | sd_delta |
|---|---|---|---|---|---|---|

| | | | | | | |
|---|---|---|---|---|---|---|
| ERR | AS_TF | 5 | -27.0853 | 6.597098 | -24.3954 | 8.169975 |
| ERR | TF | 11 | -18.8675 | 13.13358 | -19.0035 | 8.784958 |
| ERR | BAFF | 5 | -23.1721 | 5.08512 | -21.705 | 10.49697 |
| ERR | AS | 29 | -27.2676 | 13.80095 | -26.1478 | 13.10546 |
| HHRR | AS_TF | 5 | -1.9335 | 0.603855 | -1.57273 | 0.904865 |
| HHRR | TF | 11 | -2.10853 | 1.266618 | -1.97402 | 1.218765 |
| HHRR | BAFF | 5 | -1.92826 | 0.924928 | -2.21477 | 0.628891 |
| HHRR | AS | 29 | -1.44866 | 0.965574 | -1.50464 | 0.832289 |

S13. Treatment type model performance

| AIC | AICc | BIC | R2_conditional | R2_marginal | ICC | RMSE | Sigma | ScoreType |
|---|---|---|---|---|---|---|---|---|
| 681.6084 | 684.0803 | 707.6601 | 0.754186 | 0.668278 | 0.258975 | 7.181364 | 7.65527 | ERR |

| | | | | | | | | |
|---|---|---|---|---|---|---|---|---|
| 215.1493 | 217.6212 | 241.201 | 0.705649 | 0.621256 | 0.222823 | 0.572096 | 0.609268 | HHRR |

S14. Physicochemical mixed models

| effect | term | estimate | std.error | statistic | df | p.value | conf.low | conf.high | R2_marginal | R2_conditional | is_singular | formula_used | diagnostics_flagged | PhysChem_Metric | ScoreType | model | p.adj |
|---|---|---|---|---|---|---|---|---|---|---|---|---|---|---|---|---|---|
| fixed | Value_log | -4.1501 | 3.093557 | -1.34153 | 93.44384 | 0.182999 | -10.2929 | 1.992702 | 0.635177 | 0.744368 | FALSE | simplified | TRUE | pH | ERR | lmer (simplified random effects) | 0.773404 |
| fixed | Value_log | -4.77935 | 3.652947 | -1.30835 | 93.86664 | 0.193948 | -12.0325 | 2.473799 | 0.633494 | 0.745225 | FALSE | simplified | TRUE | Alkalinity_as_CaCO3_(mg/l) | ERR | lmer (simplified random effects) | 0.773404 |

| | | | | | | | | | | | | | | | | | |
|---|---|---|---|---|---|---|---|---|---|---|---|---|---|---|---|---|---|
| fixed | Value_log | 0.818732 | 1.032256 | 0.793148 | 94.96608 | 0.429669 | -1.23057 | 2.86803 | 0.633818 | 0.737615 | FALSE | simplified | TRUE | Ammoniacal_Nitrogen_as_N_(mg/l) | ERR | lmer (simplified random effects) | 0.773404 |
| fixed | Value_log | 2.961477 | 1.967902 | 1.504891 | 94.20013 | 0.135698 | -0.94573 | 6.868684 | 0.646209 | 0.736541 | FALSE | simplified | TRUE | Biochemical_Oxygen_Demand_Total_(mg/l) | ERR | lmer (simplified random effects) | 0.773404 |
| fixed | Value_log | 3.065499 | 3.2832 | 0.933692 | 94.96758 | 0.352831 | -3.45251 | 9.583504 | 0.637693 | 0.736141 | FALSE | simplified | TRUE | Chemical_Oxygen_Demand_Total_(mg/l) | ERR | lmer (simplified random effects) | 0.773404 |

| | | | | | | | | | | | | | | | | | |
|---|---|---|---|---|---|---|---|---|---|---|---|---|---|---|---|---|---|
| fixed | Value_log | 0.308398 | 0.941332 | 0.327619 | 94.79516 | 0.743922 | -1.56043 | 2.17723 | 0.628817 | 0.734108 | FALSE | simplified | TRUE | Dissolved_ammonium_(NH4)_(mg/l) | ERR | lmer (simplified random effects) | 0.810049 |
| fixed | Value_log | 0.415741 | 0.515202 | 0.806948 | 90.20099 | 0.421819 | -0.60777 | 1.439249 | 0.631646 | 0.734806 | FALSE | simplified | TRUE | Dissolved_chloride_(mg_Cl/L) | ERR | lmer (simplified random effects) | 0.773404 |
| fixed | Value_log | -0.24105 | 0.804117 | -0.29976 | 90.33541 | 0.765046 | -1.83848 | 1.356393 | 0.626832 | 0.735406 | FALSE | simplified | TRUE | Dissolved_fluoride_(mg_F/L) | ERR | lmer (simplified random effects) | 0.810049 |

| | | | | | | | | | | | | | | | | | |
|---|---|---|---|---|---|---|---|---|---|---|---|---|---|---|---|---|---|
| fixed | Value_log | 0.33833 | 0.617256 | 0.548119 | 77.9917 | 0.585175 | -0.89053 | 1.567193 | 0.666847 | 0.769594 | FALSE | simplified | TRUE | Dissolved_nitrate_(NO3) | ERR | lmer (simplified random effects) | 0.810049 |
| fixed | Value_log | -0.17295 | 0.438848 | -0.39411 | 90.22959 | 0.694432 | -1.04477 | 0.698866 | 0.627873 | 0.734968 | FALSE | simplified | TRUE | Dissolved_nitrite_(mg_NO2/L) | ERR | lmer (simplified random effects) | 0.810049 |
| fixed | Value_log | -4.03699 | 3.339359 | -1.20891 | 92.53707 | 0.229777 | -10.6687 | 2.594752 | 0.61947 | 0.744038 | FALSE | simplified | TRUE | Dissolved_organic_carbon_(mg/L) | ERR | lmer (simplified random effects) | 0.773404 |

| | | | | | | | | | | | | | | | | | |
|---|---|---|---|---|---|---|---|---|---|---|---|---|---|---|---|---|---|
| fixed | Value_log | 4.867228 | 2.916136 | 1.669067 | 79.80907 | 0.099022 | -0.93628 | 10.67074 | 0.634483 | 0.744783 | FALSE | simplified | TRUE | Dissolved_sulphate_(mg_SO4/L) | ERR | lmer (simplified random effects) | 0.773404 |
| fixed | Value_log | -1.72876 | 1.660742 | -1.04095 | 94.95176 | 0.30054 | -5.02577 | 1.568254 | 0.617986 | 0.746521 | FALSE | simplified | TRUE | Soluble_reactive_phosphorus_(?g/L) | ERR | lmer (simplified random effects) | 0.773404 |
| fixed | Value_log | 0.130807 | 2.163736 | 0.060454 | 92.84414 | 0.951924 | -4.16604 | 4.427654 | 0.628084 | 0.738662 | FALSE | simplified | TRUE | Suspended_Solids_(mg/l) | ERR | lmer (simplified random effects) | 0.951924 |

| | | | | | | | | | | | | | | | | | |
|---|---|---|---|---|---|---|---|---|---|---|---|---|---|---|---|---|---|
| fixed | Value_log | 0.647259 | 2.055873 | 0.314834 | 95.47128 | 0.753575 | -3.4339 | 4.728422 | 0.629338 | 0.737646 | FALSE | simplified | TRUE | Suspended_solids_mg/L | ERR | lmer (simplified random effects) | 0.810049 |
| fixed | Value_log | 4.073911 | 3.926737 | 1.03748 | 89.98839 | 0.302292 | -3.72725 | 11.87507 | 0.632458 | 0.735216 | FALSE | simplified | TRUE | Total_dissolved_nitrogen_(mg_N/L) | ERR | lmer (simplified random effects) | 0.773404 |
| fixed | Value_log | -1.0121 | 2.165363 | -0.46741 | 94.9463 | 0.641281 | -5.31092 | 3.286718 | 0.621165 | 0.740032 | FALSE | simplified | TRUE | Total_dissolved_phosphorus_(?g/L) | ERR | lmer (simplified random effects) | 0.810049 |

| | | | | | | | | | | | | | | | | | |
|---|---|---|---|---|---|---|---|---|---|---|---|---|---|---|---|---|---|
| fixed | Value_log | 1.313154 | 2.170823 | 0.604911 | 95.38363 | 0.546675 | -2.99625 | 5.622558 | 0.633999 | 0.735751 | FALSE | simplified | TRUE | Total_phosphorus_(?g/L) | ERR | lmer (simplified random effects) | 0.810049 |
| fixed | Value_log | -0.35623 | 0.240835 | -1.47916 | 88.84799 | 0.142635 | -0.83478 | 0.122312 | 0.604255 | 0.693456 | FALSE | simplified | TRUE | pH | HHRR | lmer (simplified random effects) | 0.513486 |
| fixed | Value_log | 0.098024 | 0.288487 | 0.339785 | 91.59416 | 0.734796 | -0.47497 | 0.671018 | 0.587219 | 0.692871 | FALSE | simplified | TRUE | Alkalinity_as_CaCO3_(mg/l) | HHRR | lmer (simplified random effects) | 0.77802 |

| | | | | | | | | | | | | | | | | | |
|---|---|---|---|---|---|---|---|---|---|---|---|---|---|---|---|---|---|
| fixed | Value_log | -0.04374 | 0.081315 | -0.53796 | 94.98779 | 0.591864 | -0.20517 | 0.117686 | 0.588135 | 0.693618 | FALSE | simplified | TRUE | Ammoniacal_Nitrogen_as_N_(mg/l) | HHRR | lmer (simplified random effects) | 0.725967 |
| fixed | Value_log | 0.282322 | 0.154349 | 1.82911 | 94.95602 | 0.070522 | -0.0241 | 0.588746 | 0.615011 | 0.687247 | FALSE | simplified | TRUE | Biochemical_Oxygen_Demand_Total_(mg/l) | HHRR | lmer (simplified random effects) | 0.42313 |
| fixed | Value_log | 0.119458 | 0.258691 | 0.461778 | 94.30315 | 0.645304 | -0.39416 | 0.633074 | 0.594909 | 0.686343 | FALSE | simplified | TRUE | Chemical_Oxygen_Demand_Total_(mg/l) | HHRR | lmer (simplified random effects) | 0.725967 |

| | | | | | | | | | | | | | | | | | |
|---|---|---|---|---|---|---|---|---|---|---|---|---|---|---|---|---|---|
| fixed | Value_log | 0.048743 | 0.073978 | 0.658882 | 94.8859 | 0.511567 | -0.09812 | 0.195611 | 0.596228 | 0.682868 | FALSE | simplified | TRUE | Dissolved_ammonium_(NH4)_(mg/l) | HHRR | lmer (simplified random effects) | 0.725967 |
| fixed | Value_log | 0.115994 | 0.039226 | 2.957077 | 91.0163 | 0.003956 | 0.038077 | 0.193912 | 0.625323 | 0.707938 | FALSE | simplified | TRUE | Dissolved_chloride_(mg_Cl/L) | HHRR | lmer (simplified random effects) | 0.071205 |
| fixed | Value_log | -0.10556 | 0.062773 | -1.68169 | 90.90609 | 0.096061 | -0.23026 | 0.019128 | 0.598601 | 0.69601 | FALSE | simplified | TRUE | Dissolved_fluoride_(mg_F/L) | HHRR | lmer (simplified random effects) | 0.432277 |

| | | | | | | | | | | | | | | | | | |
|---|---|---|---|---|---|---|---|---|---|---|---|---|---|---|---|---|---|
| fixed | Value_log | 0.022661 | 0.048927 | 0.463166 | 78.76096 | 0.644523 | -0.07473 | 0.120052 | 0.625139 | 0.715123 | FALSE | simplified | TRUE | Dissolved_nitrate_(NO3) | HHRR | lmer (simplified random effects) | 0.725967 |
| fixed | Value_log | -0.07051 | 0.034033 | -2.07193 | 90.89076 | 0.041106 | -0.13812 | -0.00291 | 0.605763 | 0.698707 | FALSE | simplified | TRUE | Dissolved_nitrite_(mg_NO2/L) | HHRR | lmer (simplified random effects) | 0.36995 |
| fixed | Value_log | -0.24616 | 0.264318 | -0.93128 | 93.47894 | 0.354106 | -0.771 | 0.278693 | 0.582707 | 0.691187 | FALSE | simplified | TRUE | Dissolved_organic_carbon_(mg/L) | HHRR | lmer (simplified random effects) | 0.708212 |

| | | | | | | | | | | | | | | | | |
|---|---|---|---|---|---|---|---|---|---|---|---|---|---|---|---|---|
| fixed | Value_log | 0.24193 | 0.226555 | 1.067864 | 68.76815 | 0.289316 | -0.21006 | 0.693923 | 0.592857 | 0.687036 | FALSE | simplified | TRUE | Dissolved_sulphate_(mg_SO4/L) | HHRR | lmer (simplified random effects) | 0.705803 |
| fixed | Value_log | 0.095686 | 0.130403 | 0.733774 | 92.24308 | 0.464947 | -0.1633 | 0.354669 | 0.599656 | 0.680263 | FALSE | simplified | TRUE | Soluble_reactive_phosphorus_(?g/L) | HHRR | lmer (simplified random effects) | 0.725967 |
| fixed | Value_log | 0.107103 | 0.170485 | 0.628224 | 93.73613 | 0.531385 | -0.23141 | 0.445617 | 0.591581 | 0.690918 | FALSE | simplified | TRUE | Suspended_Solids_(mg/l) | HHRR | lmer (simplified random effects) | 0.725967 |

| | | | | | | | | | | | | | | | | | |
|---|---|---|---|---|---|---|---|---|---|---|---|---|---|---|---|---|---|
| fixed | Value_log | -0.11987 | 0.162198 | -0.73902 | 95.97243 | 0.461696 | -0.44183 | 0.202094 | 0.590337 | 0.685943 | FALSE | simplified | TRUE | Suspended_solids_mg/L | HHRR | lmer (simplified random effects) | 0.725967 |
| fixed | Value_log | 0.36827 | 0.303481 | 1.213489 | 80.91049 | 0.228474 | -0.23557 | 0.972112 | 0.602543 | 0.679675 | FALSE | simplified | TRUE | Total_dissolved_nitrogen_(mg_N/L) | HHRR | lmer (simplified random effects) | 0.685421 |
| fixed | Value_log | 0.171227 | 0.169023 | 1.013042 | 92.23486 | 0.31369 | -0.16446 | 0.50691 | 0.605282 | 0.677941 | FALSE | simplified | TRUE | Total_dissolved_phosphorus_(?g/L) | HHRR | lmer (simplified random effects) | 0.705803 |

| | | | | | | | | | | | | | | | | | |
|---|---|---|---|---|---|---|---|---|---|---|---|---|---|---|---|---|---|
| fixed | Value_log | -0.03259 | 0.171909 | -0.18956 | 95.82654 | 0.850053 | -0.37383 | 0.308657 | 0.586785 | 0.685457 | FALSE | simplified | TRUE | Total_phosphorus_(?g/L) | HHRR | lmer (simplified random effects) | 0.850053 |

S15. Censoring frequency

| PhysChem_Metric | n_total | n_censored | pct_censored |
|---|---|---|---|
| pH | 203 | 0 | 0 |
| Alkalinity_as_CaCO3_(mg/l) | 203 | 0 | 0 |
| Ammoniacal_Nitrogen_as_N_(mg/l) | 203 | 14 | 6.9 |

| | | | |
|---|---|---|---|
| Biochemical_Oxygen_Demand_Total_(mg/l) | 203 | 8 | 3.9 |
| Chemical_Oxygen_Demand_Total_(mg/l) | 203 | 2 | 1 |
| Dissolved_ammonium_(NH4)_(mg/l) | 203 | 0 | 0 |
| Dissolved_chloride_(mg_Cl/L) | 203 | 0 | 0 |
| Dissolved_fluoride_(mg_F/L) | 203 | 0 | 0 |

| | | | |
|---|---|---|---|
| Dissolved_nitrate_(NO3) | 178 | 0 | 0 |
| Dissolved_nitrite_(mg_NO2/L) | 203 | 0 | 0 |
| Dissolved_organic_carbon_(mg/L) | 201 | 0 | 0 |
| Dissolved_sulphate_(mg_SO4/L) | 201 | 0 | 0 |
| Soluble_reactive_phosphorus_(?g/L) | 203 | 0 | 0 |
| Suspended_Solids_(mg/l) | 203 | 22 | 10.8 |

| | | | |
|---|---|---|---|
| Suspended_solids_mg/L | 205 | 0 | 0 |
| Total_dissolved_nitrogen_(mg_N/L) | 201 | 0 | 0 |
| Total_dissolved_phosphorus_(?g/L) | 203 | 0 | 0 |
| Total_phosphorus_(?g/L) | 205 | 0 | 0 |

S16. **Assembly quality comparison between influent and effluent metagenomes**

| Metric | Mann-Whitney U | p-value |
|---|---|---|

| | | |
|---|---|---|
| N50 | 1515.5 | 0.068 |
| Number of contigs | 1178 | 0.622 |
| Total assembled bases | 1339 | 0.542 |

**S17. N50 correlations with MetaCompare risk scores**

| Risk metric | Spearman rho | p-value |
|---|---|---|
| ERR | 0.436 | $5.91 \times 10^{-6}$ |
| HHRR | 0.352 | $3.29 \times 10^{-4}$ |

**S18. Effect of treatment stage after accounting for assembly quality**

| Outcome | Predictor | Estimate | SE | p-value | 95% CI |
|---|---|---|---|---|---|
| ERR | Final effluent | -23.67 | 1.62 | <0.001 | -26.88 to -20.47 |
| ERR | N50 | 3.51 | 0.68 | <0.001 | 2.16 to 4.85 |
| HHRR | Final effluent | -1.67 | 0.14 | <0.001 | -1.94 to -1.41 |
| HHRR | N50 | 0.15 | 0.06 | 0.012 | 0.03 to 0.26 |
